# Diagnostic Value of Elastography in Differentiating Parathyroid Adenoma from Hyperplasia: A Systematic Review and Meta-Analysis

**DOI:** 10.64898/2025.12.11.25342045

**Authors:** Amir Hassankhani, Payam Jannatdoust, Parya Valizadeh, Melika Amoukhteh, Abbas Mohammadi, Ali Gholamrezanezhad, Ali Haq

**Affiliations:** Department of Graduate Medical Education, Valley Health System, Las Vegas, NV, USA; School of Medicine, Tehran University of Medical Sciences, Tehran, Iran; Department of Internal Medicine, Valley Health System, Las Vegas, NV, USA; Department of Radiology, Cedars-Sinai Medical Center, Los Angeles, CA, USA

**Keywords:** Parathyroid Neoplasms, Primary, Parathyroid Hyperplasia, Ultrasonography, Elastography, Diagnostic Accuracy, Sensitivity and Specificity

## Abstract

**Background:** Differentiating parathyroid adenoma from hyperplasia is critical for surgical planning, but conventional imaging often cannot reliably distinguish these lesions. Ultrasound elastography offers quantitative assessment of tissue stiffness and may improve preoperative characterization.

**Purpose:** To evaluate the diagnostic accuracy of ultrasound elastography in differentiating parathyroid adenoma from hyperplasia.

**Methods:** A systematic review and meta-analysis was conducted in accordance with PRISMA guidelines. PubMed, Scopus, and Embase were searched through July 2025, for studies assessing elastography for parathyroid lesion differentiation. Data on sensitivity, specificity, and other diagnostic metrics were extracted and pooled using a bivariate random-effects model in R software.

**Results:** Five studies comprising 579 parathyroid lesions were included, of which four studies with 352 adenomas and 202 hyperplasias were eligible for pooled analysis. Pooled sensitivity and specificity of elastography were 83.3% (95% CI: 74.3–89.6%) and 79.1% (95% CI: 65.5–88.3%), respectively, with an area under the SROC curve of 0.88 (95% CI: 0.79–0.92). Pooled likelihood ratios were 4.14 for a positive test and 0.216 for a negative test. Fagan nomogram analysis showed that, for a patient with a 50% pre-test probability of adenoma, a positive result increased post-test probability to 81%, while a negative result decreased it to 18%.

**Conclusion:** Ultrasound elastography demonstrates good diagnostic performance in distinguishing parathyroid adenomas from hyperplasia and may help inform preoperative planning.

## Introduction

Differentiating parathyroid adenoma from hyperplasia is essential, as surgical strategies differ: a single-gland adenoma can often be cured with targeted excision, whereas hyperplasia typically requires subtotal or total parathyroidectomy. Accurate preoperative characterization is therefore critical to avoid persistent disease or reoperation (1–4).

High-resolution ultrasonography and technetium-99m sestamibi scintigraphy remain standard for localizing abnormal parathyroid tissue (5–7). While effective for gland detection, these modalities may have limited ability to distinguish adenomas from hyperplastic glands or to differentiate parathyroid lesions from adjacent cervical structures such as thyroid nodules and lymph nodes. This limitation is particularly relevant for minimally invasive parathyroidectomy, where precise localization directly impacts surgical success and morbidity (7–9).

Ultrasound elastography offers a promising complementary approach by quantitatively assessing tissue stiffness (10–12). Parathyroid adenomas generally appear stiffer due to compact cellular architecture, reduced adipose content, and thickened capsules, whereas hyperplastic glands exhibit more heterogeneous elasticity (12, 13). By providing objective biomechanical data, elastography has the potential to improve differentiation between adenoma and hyperplasia and enhance preoperative planning (12–14). Although early reports are encouraging, wide variability in diagnostic accuracy has limited clinical adoption. To address this gap, we conducted a systematic review and meta-analysis to evaluate the diagnostic performance of elastography in distinguishing parathyroid adenoma from hyperplasia, clarifying its role as a radiologic adjunct in preoperative assessment and decision-making.

## Methods

### Study Design

This systematic review and meta-analysis were conducted in accordance with the Preferred Reporting Items for Systematic Reviews and Meta-Analyses (PRISMA) guidelines (15).

### Search Strategy

A comprehensive literature search was performed in PubMed, Embase, and Scopus through July 17, 2025, to identify studies evaluating elastography for differentiating parathyroid adenoma from hyperplasia. Search terms included combinations of keywords and vocabulary related to elastography, parathyroid lesions, adenoma, hyperplasia, and diagnostic accuracy. The general search strategy was as follows: (“ultrasound elastography” OR “shear wave elastography” OR “strain elastography” OR SWE OR USE) AND (“parathyroid adenoma” OR “parathyroid hyperplasia” OR “parathyroid lesion*”) AND (“diagnostic accuracy” OR sensitivity OR specificity OR “receiver operating characteristic” OR ROC OR AUC OR detect* OR diagnos*).

Search terms were adapted to each database, and all results were imported into reference management software for deduplication. Reference lists of included articles and relevant reviews were manually screened to identify additional eligible studies.

### Eligibility Criteria

Inclusion criteria:

- Human participants with suspected or confirmed parathyroid lesions.
- Elastography performed to assess parathyroid adenomas and hyperplasia.
- Diagnostic performance metrics reported, or sufficient data to calculate sensitivity and specificity.
- Histopathology, surgical findings, or validated clinical/imaging follow-up used as the reference standard.

Exclusion criteria:

- Studies evaluating only one lesion type without differentiation.
- Studies limited to non-parathyroid neck lesions.
- Case reports, small case series (<5 patients), conference abstracts without full data, reviews, or editorials.
- Non-English publications without translation.

### Study Selection

Two independent reviewers screened titles and abstracts for relevance. Full texts of potentially eligible studies were retrieved and assessed against inclusion criteria. Discrepancies were resolved by discussion or by a third reviewer.

### Data Extraction

Two reviewers independently extracted:

- Author, year, country.
- Study design and sample size.
- Patient demographics and inclusion criteria.
- Reference standard.
- Elastography technique and parameters.
- Diagnostic metrics (sensitivity, specificity, true positives, false positives, true negatives, false negatives).

Discrepancies were resolved by consensus.

### Quality Assessment

Methodological quality was assessed with the Quality Assessment of Diagnostic Accuracy Studies-2 (QUADAS-2). Risk of bias and applicability were evaluated across patient selection, index test, reference standard, and flow/timing (16). Disagreements were resolved through discussion.

### Statistical Analysis

Diagnostic accuracy meta-analysis was conducted using 2×2 contingency tables from each study. A bivariate random-effects model (17) was applied to estimate pooled sensitivity and specificity. Summary receiver operating characteristic (SROC) curves were generated, and the area under the curve (AUC) was estimated via parametric bootstrapping with 2000 replications (18).

Between-study heterogeneity was assessed with the I² statistic. Values >50% indicated notable variability (19).

Fagan nomograms were constructed at pre-test probabilities of 25%, 50%, and 75% to illustrate clinical impact.

Analyses were conducted in R (version 4.2.1; R Foundation for Statistical Computing, Vienna, Austria) using the “meta,” “mada” (20), “dmetatools,” and “metafor” (21) packages.

## Results

### Study Selection and Screening Process

The search yielded 1,441 records. After removal of duplicates, 862 unique articles remained. Following title and abstract screening, 847 studies were excluded. Full texts of 15 potentially relevant articles were reviewed, of which 10 were excluded for not meeting inclusion criteria (failure to differentiate adenoma from hyperplasia or lack of diagnostic accuracy data).

Ultimately, 5 studies met all eligibility requirements and were included in the final analysis. The study selection process is outlined in the PRISMA flow diagram (Figure 1).

**Figure 1.**
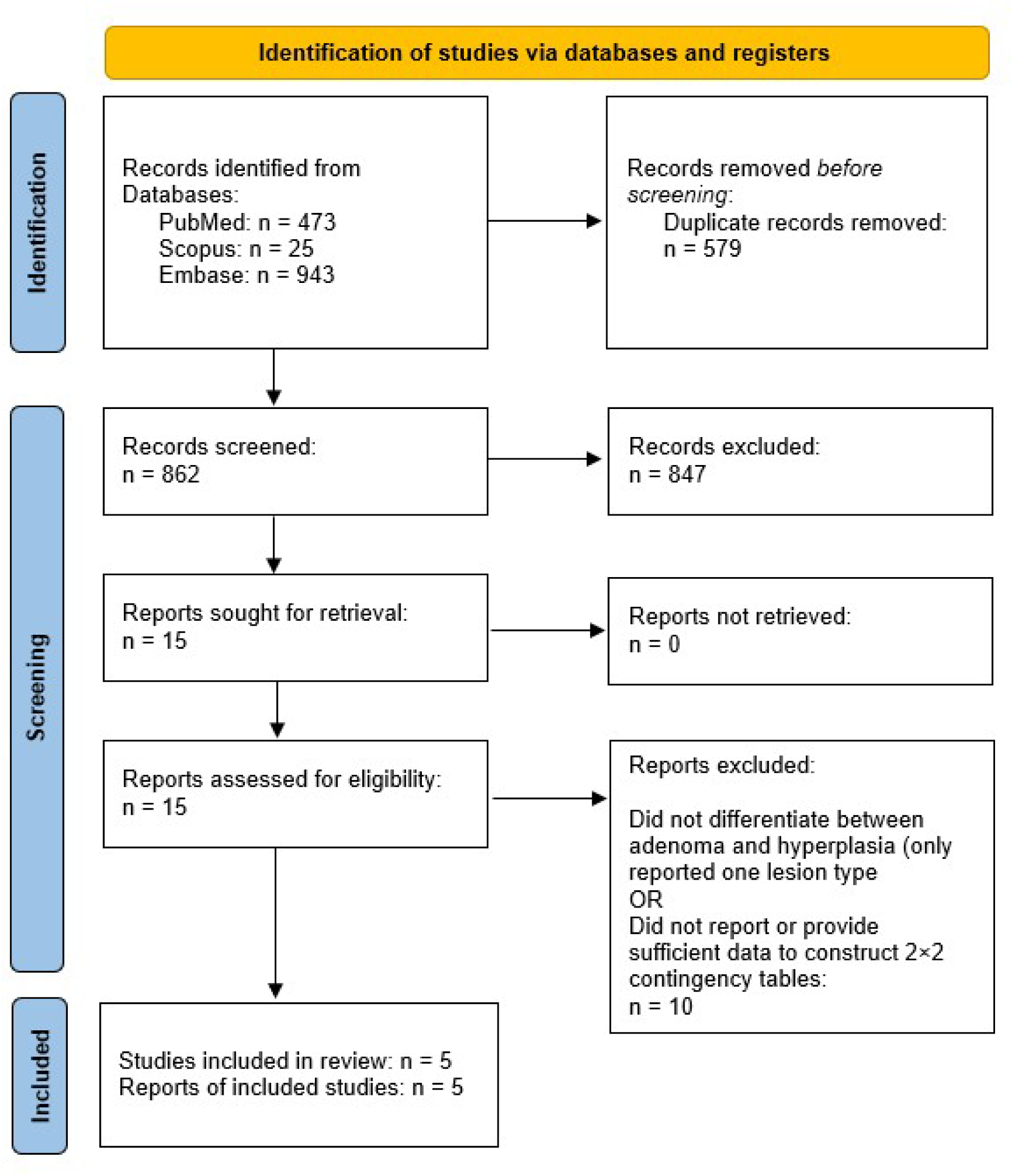
PRISMA 2020 flow diagram summarizing the study selection process for inclusion in the meta-analysis.

### Study Characteristics

The five included studies encompassed 579 parathyroid lesions: 352 adenomas, 156 hyperplasias, and 25 atypical adenomas. All were prospective, single-center studies published between 2009 and 2022. Final diagnoses were histopathologically confirmed in most cases; in secondary hyperparathyroidism, clinical and biochemical criteria were also used.

Elastography methods included strain elastography, two-dimensional shear wave elastography (2D-SWE), and Virtual Touch tissue imaging quantification (VTIQ). Diagnostic indices included strain ratio, shear wave velocity, and elastic modulus. One study (13), including 25 atypical adenomas, did not report diagnostic thresholds and was excluded from pooled quantitative analysis (Table 1).

**Table 1.**
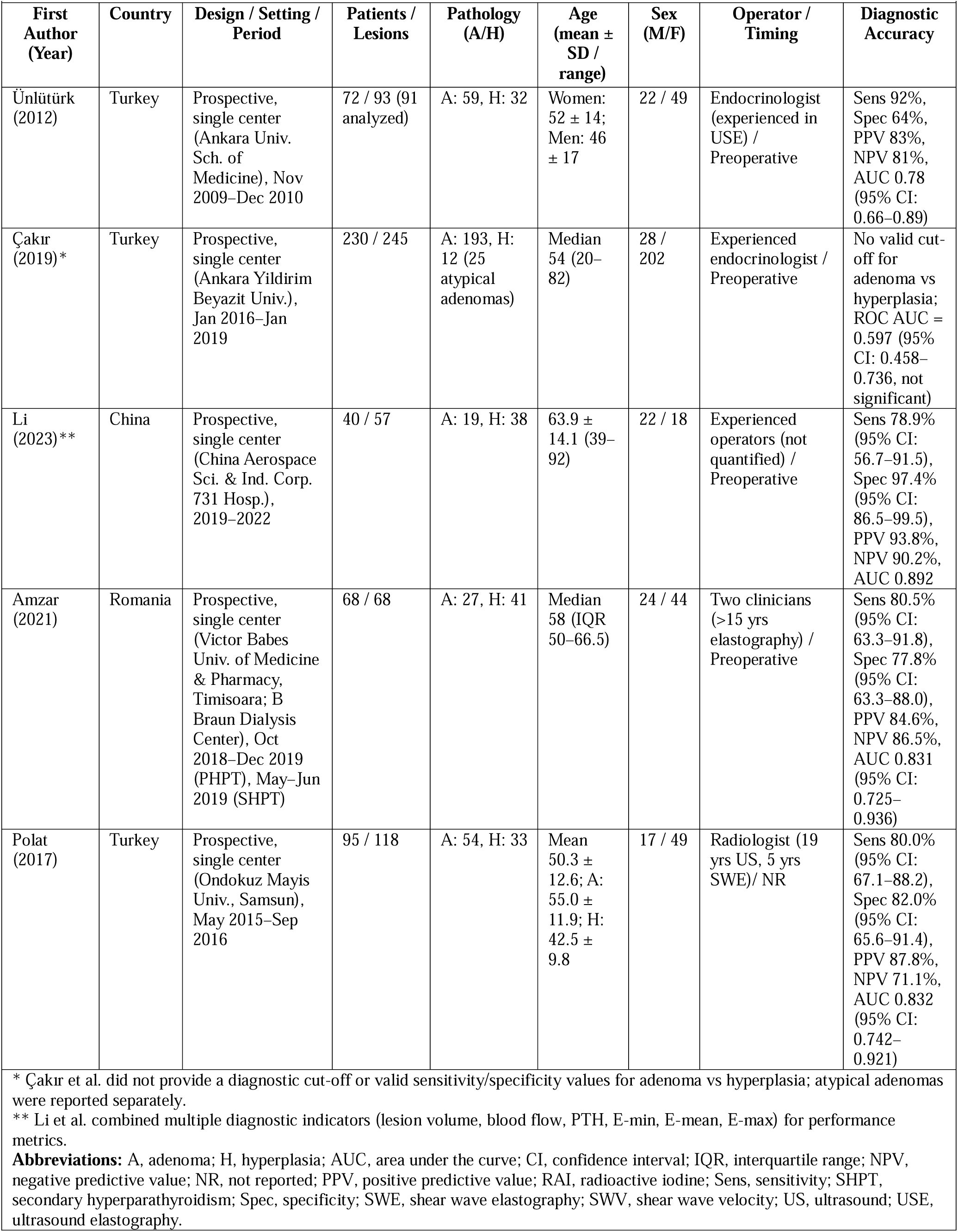
Study characteristics and diagnostic performance of elastography for differentiating parathyroid adenomas from hyperplasia.

### Quality Assessment

QUADAS-2 results are summarized in Table 2. Most studies demonstrated low or moderate risk of bias. Applicability concerns were generally low, with the exception of Amzar (12), where moderate concern was noted for the reference standard.

**Table 2.** Quality assessment of included studies using the QUADAS-2 tool.

| First Author, Year | Risk of Bias |  |  |  | Applicability Concerns |  |  |
| --- | --- | --- | --- | --- | --- | --- | --- |
|  | Patient Selection | Index Test | Reference Standard | Flow and Timing | Patient Selection | Index Test | Reference Standard |
| Ünlü et al., 2012 | Low | Low | Low | Low | Low | Low | Low |
| Li, 2023 | Unclear | Low | Low | Moderate | Low | Low | Low |
| Cakir, 2019 | Low | Low | Low | Low | Low | Low | Low |
| Amzar, 2021 | Moderate | Low | Moderate | Low | Low | Low | Moderate |
| Polat, 2017 | Low | Moderate | Low | Low | Low | Low | Low |

QUADAS-2, Quality Assessment of Diagnostic Accuracy Studies-2

### Meta-Analysis

Pooled sensitivity was 83.3% (95% CI: 74.3–89.6) and specificity 79.1% (95% CI: 65.5–88.3), with no statistical heterogeneity detected (I² = 0.0%) (Figure 2). The SROC analysis yielded an AUC of 0.88 (95% CI: 0.79–0.92) (Figure 3).

**Figure 2.**
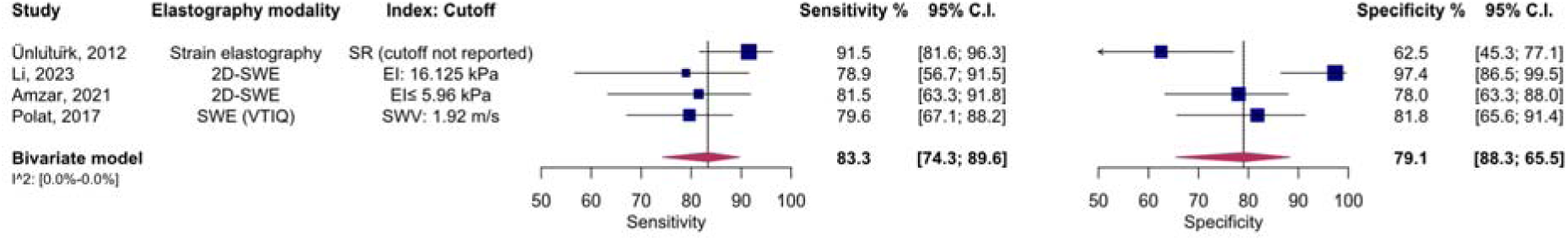
Forest plot of individual studies and pooled estimates of elastography for differentiating parathyroid adenoma from parathyroid hyperplasia. 2D-SWE, two-dimensional shear wave elastography; CI, confidence interval; EI, elasticity index; SR, strain ratio; SWE, shear wave elastography; SWV, shear wave velocity; VTIQ, Virtual Touch tissue imaging quantification.

**Figure 3.**
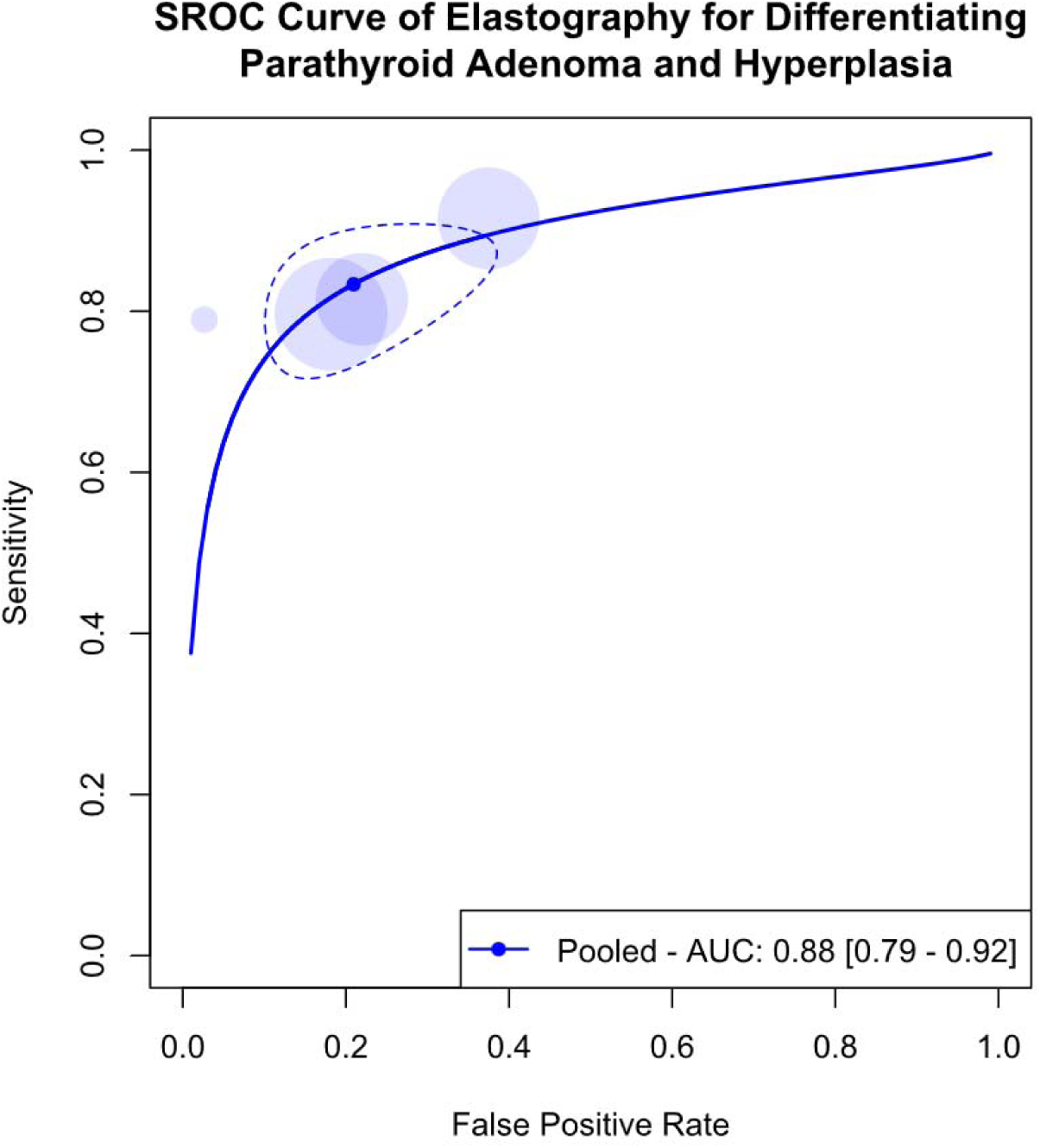
SROC curve of elastography for differentiating parathyroid adenoma from parathyroid hyperplasia, along with the pooled area under the curve (AUC). AUC, area under the curve; SROC, summary receiver operating characteristic.

Fagan nomograms demonstrated that at a pre-test probability of 50%, a positive elastography result increased the probability of adenoma to 80.5%, while a negative result reduced it to 17.8%. Corresponding post-test probabilities were 58.0% and 6.7% at a 25% pre-test probability, and 92.6% and 39.3% at a 75% pre-test probability. The pooled likelihood ratios were 4.14 for a positive test and 0.216 for a negative test (Figure 4).

**Figure 4.**
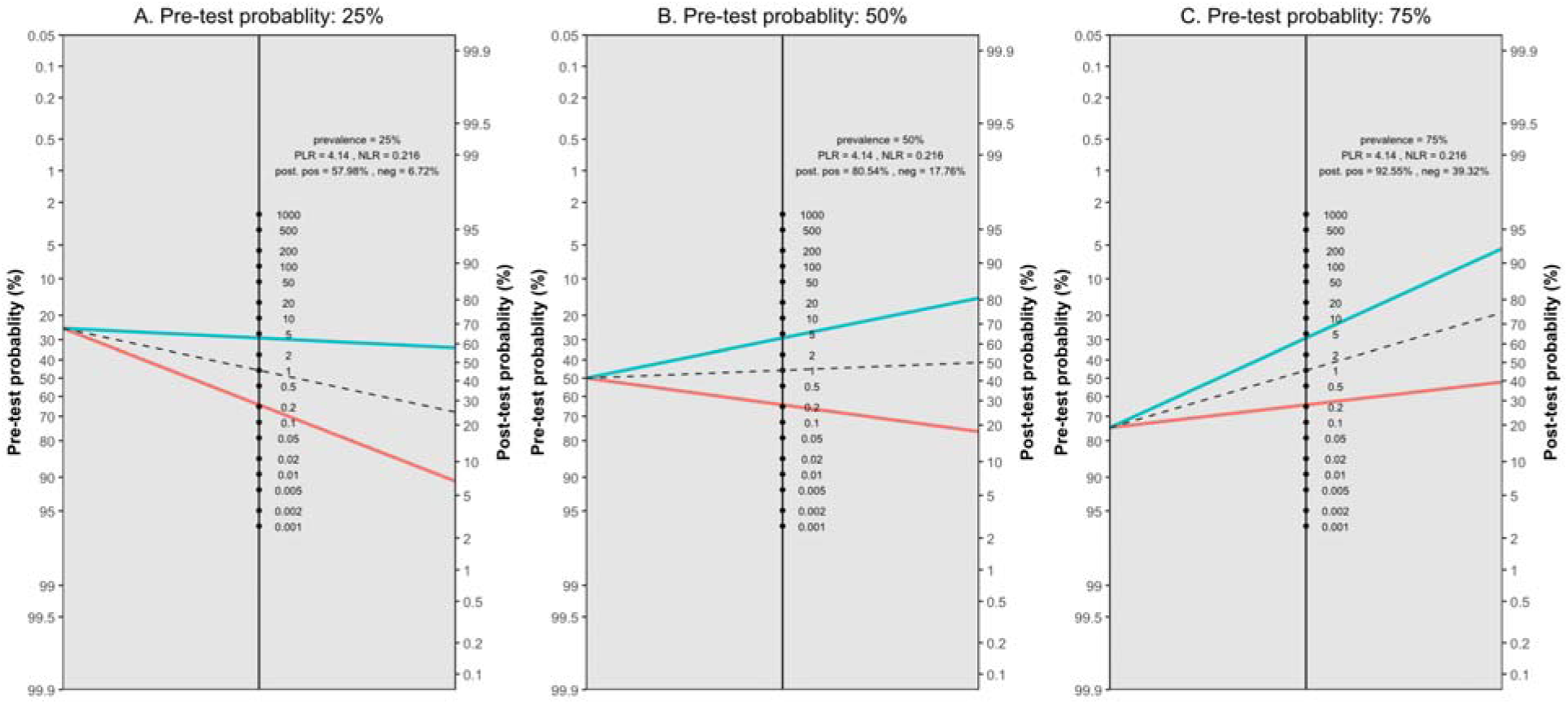
Fagan nomograms assessing the clinical utility of elastography for differentiating parathyroid adenoma from parathyroid hyperplasia at pre-test probabilities of 25% (A), 50% (B), and 75% (C). NLR, negative likelihood ratio; PLR, positive likelihood ratio.

## Discussion

In this systematic review and meta-analysis, elastography showed good overall diagnostic performance in distinguishing parathyroid adenomas from hyperplasia. These findings suggest meaningful clinical utility for elastography in the preoperative assessment of parathyroid lesions.

The relatively high sensitivity observed implies that elastography can reliably detect adenomas, which is particularly important in planning minimally invasive parathyroidectomy. The underlying biological rationale is consistent with prior observations: adenomas are characterized by firm, homogeneous tissue resulting from compact cells and limited adipose tissue, while hyperplasia is associated with softer, more irregular tissue mechanics (22–24). Elastography directly quantifies these tissue stiffness differences, which likely accounts for its diagnostic value (11, 22, 25).

Specificity was also favorable, suggesting that elastography can correctly identify a majority of hyperplastic glands. Nevertheless, some overlap in stiffness values was observed. This is likely attributable to histological variability, as hyperplastic glands may occasionally demonstrate increased stiffness due to fibrosis or nodular change, while some adenomas appear softer if they contain adipose or cystic components (26, 27). Such overlap highlights the importance of integrating elastography with other imaging modalities rather than relying on it in isolation. Advanced methods such as 2D-SWE and VTIQ may reduce misclassification (24, 28–30), though standardization of technique and diagnostic thresholds remains necessary.

The Fagan nomogram analysis highlights the clinical impact of elastography. In a patient with a 50% pre-test probability of adenoma, a positive elastography result raised the post-test probability to approximately 81%, while a negative result reduced it to 18%. These shifts in probability are substantial enough to influence surgical planning, but they fall short of providing definitive diagnostic certainty. Therefore, elastography is best applied in combination with conventional imaging such as high-resolution ultrasonography and sestamibi scintigraphy, optimizing both localization and lesion characterization.

In our analysis, although statistical heterogeneity was not detected, methodological variability between studies should be acknowledged. Differences in elastography techniques, measurement parameters, cut-off values, and operator expertise may affect diagnostic performance and limit generalizability. Furthermore, the relatively small number of included studies prevented subgroup or meta-regression analyses that might clarify the impact of factors such as lesion size, patient comorbidities, or differences between primary and secondary hyperparathyroidism.

Despite these limitations, this study has notable strengths. The use of histopathology or validated clinical criteria as reference standards minimized misclassification bias and enhanced reliability of pooled estimates. The inclusion of studies from multiple geographic regions also broadens the relevance of the findings.

Future research should focus on multicenter prospective trials with larger cohorts, standardized acquisition protocols, and uniform diagnostic thresholds. Incorporating elastography into multimodal diagnostic algorithms alongside clinical, biochemical, and imaging parameters may further improve diagnostic accuracy. In addition, artificial intelligence–based image analysis offers the potential for automated and reproducible stiffness measurements, which could mitigate operator dependence and enhance diagnostic precision.

## Conclusion

This systematic review and meta-analysis demonstrate that ultrasound elastography provides good diagnostic performance in differentiating parathyroid adenomas from hyperplasia. While not definitive as a standalone test, elastography serves as a valuable complementary modality. Integration into multimodal diagnostic workflows can improve surgical planning and may reduce reoperation risk, particularly in centers pursuing minimally invasive parathyroidectomy.

## Data Availability Statement

The datasets analyzed during the current study are available from the corresponding author on reasonable request.

## Declaration of Generative AI and AI-assisted technologies in the writing process

During the preparation of this work, the authors used ChatGPT to improve readability and clarity. After using this tool, the authors reviewed and edited the content as needed and take full responsibility for the content of this publication.

